# Xylazine in Drug Seizure Reports and Overdose Deaths in the US, 2018-2022

**DOI:** 10.1101/2023.08.24.23294567

**Authors:** Manuel Cano, Raminta Daniulaitye, Flavio Marsiglia

## Abstract

Xylazine is increasingly reported in street drugs and fatal overdoses in the United States (US), often in combination with synthetic opioids, yet state-level xylazine data are limited, hampering local public health responses. The present study analyzed 2018-2022 state-level data from the National Forensic Laboratory Information System (xylazine-positive reports of seized drugs analyzed by forensic laboratories), the Centers for Disease Control and Prevention (population estimates, synthetic opioid overdose mortality rates), and individual states’ medical examiner/public health agency reports (numbers of xylazine-involved overdose deaths). An ordinary least squares regression model predicted state-level synthetic opioid overdose mortality rates by xylazine seizure report rates, adjusting for US Census Region. In 2018, relatively low rates of xylazine seizure reports were observed, with 21 states reporting zero xylazine seizures. In 2022, only three states reported zero xylazine seizures, and the highest xylazine seizure report rates (per 100,000 residents) were observed in New Jersey (30.4), Rhode Island (22.7), Maryland (18.9), Virginia (15.5), New Hampshire (13.0), and Ohio (10.9). Data on 2019-2022 xylazine-involved overdose deaths were available for 21 states/DC (60 state-years), with the highest 2022 xylazine-involved overdose death rates (per 100,000 residents) in Vermont (10.5) and Connecticut (9.8). Finally, in 2021, at the state level, each additional reported xylazine seizure per 100,000 residents was associated with a 2% higher synthetic opioid overdose mortality rate (*b*=0.02, robust standard error=0.01; *p*=0.049). Overall, study results emphasize xylazine’s increasing involvement in US law enforcement drug seizure reports and overdose deaths, primarily in the East, yet also extending across the country.

---

Xylazine (a sedative/analgesic approved only for veterinary use) has been designated by the United States (US) Government as an “emerging threat” due to its increasing presence in fatal overdoses.^1^ Xylazine-involved overdose deaths in the US rose from 102 in 2018 to 3,468 in 2021,^2^ and nearly all (97%-100%) of these deaths also involved synthetic opioids such as fentanyl.^2–4^ Xylazine-involved overdose deaths are primarily concentrated in the eastern US,^2^ especially in the Northeast,^3^ yet it is less clear to what extent xylazine is involved in overdose deaths within individual states across the US, in part because xylazine-involved overdose death counts/rates are not accessible via the Centers for Disease Control and Prevention (CDC)’s national mortality database (Multiple Cause of Death via the National Vital Statistics System). *State-level* data on xylazine-involved overdose deaths are currently limited in peer-reviewed publications, including analyses from individual states (Connecticut^4^; West Virginia^5^; Tennessee^6^) and a CDC report^7^ providing ranges (e.g., 100 to 499 deaths) of fentanyl/xylazine-positive deaths for 31 states. In a 2022 publication,^8^ researchers were able to locate xylazine-involved overdose death data for only 14 jurisdictions in the US (including cities, counties, and states).

Information about xylazine’s presence in street drug supplies and in overdose deaths within individual states, cities, and counties is necessary to inform local public health agencies’ and providers’ implementation of xylazine-specific harm reduction strategies. In light of gaps in overdose death data availability and timeliness, various studies have used law enforcement drug seizure data to predict or forecast overdose deaths.^9–12^ While law enforcement drug seizures may be undertaken with the intent of reducing drug-related harms, including overdose, the “drug bust paradox” posits that fatal opioid overdoses increase after law enforcement operations such as drug seizures, as individuals who use drugs lose trusted suppliers and resume use with reduced opioid tolerance, obtaining drugs with unknown composition and potency.^13,14^ At the same time, numerous state- and county-level studies have conceptualized drug seizure measures as indicators of the *availability* of different substances within the illicit drug supply (rather than as law enforcement operations that directly affect overdose rates), finding increased levels of drug seizures involving fentanyl and/or fentanyl analogs associated with increased overdose mortality rates.^11,15–20^ Nonetheless, relatively less is known about *xylazine* seizures.

Therefore, the present study compiles and depicts available data on xylazine involvement in US drug seizure cases and overdose deaths across states, regions, and years (2018-2022). The study also hypothesizes that the extent of xylazine’s presence in states’ illicit drug supplies (as approximated via xylazine drug seizure reports) is associated with states’ levels of synthetic opioid overdose mortality, considering that xylazine is nearly always accompanied by synthetic opioids in cases of fatal overdose.^2^

## Methods

### Data Sources

#### Xylazine Seizure Data

Drug seizure data were obtained from the National Forensic Laboratory Information System (NFLIS) online Public Data Query System,^21^ which provides public-access data from 98% of US law enforcement drug seizure samples submitted to and analyzed by forensic laboratories.^22^ Results from approximately 95% of analyzed drug submissions are available in NFLIS within three months, with the remaining 5% updated as available^22^; the data used in the present study represented xylazine seizure reports for 2018-2022 recorded in the NFLIS as of July 28, 2023. (Although NFLIS drug seizure reports do not encompass the totality of drugs seized by law enforcement,^22^ in the present study, we use the term “xylazine seizure” for brevity, referring to drug seizure samples in which xylazine was detected and reported when analyzed by NFLIS-participating laboratories.) We extracted numbers of xylazine seizures for each year for each of the 50 states and District of Columbia, as well as for all states combined and for each Health and Human Services (HHS) region.

#### Overdose Mortality Data

The CDC’s national mortality database (Multiple Cause of Death from the National Vital Statistics System) represents the primary nationwide source of counts or rates of various types of overdose deaths (e.g., involving synthetic opioids, cocaine), identified via International Classification of Diseases (ICD)-10 codes. No ICD-10 code corresponds solely to xylazine,^2^ and although the ICD-10 code T42.7 (antiepileptic and sedative-hypnotic) can include both xylazine and levetiracetam,^2^ in 2021, the number of fatal drug overdoses reported in national mortality data^23^ with a T42.7 code as a “multiple [contributing] cause of death” (2,657) represented only a fraction (77%) of the total 3,468 xylazine-involved deaths in 2021 identified via a separate CDC analysis of literal text on death certificates.^2^

Since numbers of xylazine-involved overdose deaths are not available via the public-access or restricted-access version of the aforementioned CDC mortality database, we first used xylazine-involved overdose rates recently published by the CDC (based on their analysis of literal text on death certificates^2^), comprising age-adjusted rates by HHS *region* (for the year 2021, with data unavailable for three of the ten HHS regions) and for the *US overall* (for each year from 2018-2021). Since *state-level* xylazine-involved overdose counts/rates were *not* provided in this CDC publication,^2^ we next searched online for state-level counts of xylazine-involved overdose deaths for any year between 2019 and 2022 (following the example of a prior study^8^). For each state and the District of Columbia, we conducted systematic online searches by querying Google with the terms “[state name] overdose death data xylazine,” proceeding to additional search terms as necessary, including “[state name] overdose data,” “[state name] SUDORS [State Unintentional Drug Overdose Reporting System] data report,” “[state name] medical examiner overdose report,” and “[state public health agency name] overdose data.” We recorded any state-level xylazine-involved overdose death count available from an official state website (e.g., public health department website, medical examiner website), the webpage of a university/entity contracted to analyze statewide overdose data, or a peer-reviewed journal article. We recorded counts available for one-year time periods (or half of a year, which we doubled to estimate a yearly total) but not for several years combined. Considering that approximately 99% of all xylazine-involved overdose deaths also involve fentanyl,^2^ we extracted counts of xylazine-involved overdose deaths overall or overdose deaths involving both xylazine and fentanyl or opioids, as available in each data source. In cases where only the total number of overdose deaths and percentage involving xylazine was provided, we multiplied the total number of overdose deaths by the fraction involving xylazine to arrive at an estimated number of xylazine-involved overdose deaths.

Finally, we obtained 2021 state-level rates of *synthetic opioid* overdose deaths (per 100,000 population) from the Multiple Cause of Death dataset via the CDC WONDER online platform.^23^ Synthetic opioid overdose deaths were identified via an “underlying cause of death” corresponding to drug poisoning of any intent (ICD-10 codes X40-44, X60-64, X85, Y10-14) and a contributing “multiple cause of death” corresponding to ICD-10 category T40.4 (synthetic opioids excluding methadone).

#### Population Estimates

Population estimates, by state and year, as well as for the entire US and HHS regions, were obtained from the National Center for Health Statistics via the CDC WONDER online platform.^23^ These population estimates were utilized in the computation of rates of xylazine seizures and xylazine-involved overdose deaths.

### Statistical Analyses

Analyses were completed in Stata/MP 18.0 and RStudio. First, in order to visually depict regional and time patterns in rates of xylazine drug seizure reports and xylazine-involved overdose deaths, we plotted: a) a line graph depicting xylazine seizure rates and xylazine-involved overdose death rates in the US over time (2018-2021); and b) a scatter plot with each HHS region’s 2021 xylazine seizure rate (x-axis) and 2021 xylazine-involved overdose death rate (y-axis). We also computed the annual percent change in nationwide yearly rates of xylazine seizures and xylazine-involved overdose deaths for each year from 2018 to 2021. The modest number of available xylazine-involved overdose mortality rates precluded a formal analysis of correlation between xylazine seizures and xylazine-involved overdose deaths.

We next examined state-level data. To visually compare xylazine *seizure* rates between US states, we plotted the number of xylazine seizures per 100,000 residents for each state and year (2018-2022), ranking states based on their 2022 xylazine seizure rates. To examine xylazine-involved *overdose* deaths in different states, we used xylazine overdose death counts and population estimates to calculate rates per 100,000 residents (for each state and year with data available from our systematic online search). We then plotted rates of xylazine seizures and xylazine-involved overdose deaths by state and year to aid visualization of patterns over time. Nonetheless, the small number of states with xylazine mortality data again prevented any formal analysis of the association between xylazine seizures and xylazine-involved overdose deaths.

Finally, in the absence of xylazine-involved overdose mortality data for all states, we examined the extent to which states’ xylazine seizure levels may be associated with *synthetic opioid* overdose mortality, considering that nearly all xylazine-involved overdose deaths also involve synthetic opioids such as fentanyl.^2^ We used data from 2021, the most recent year with finalized (non-provisional) mortality data available for all states, in an ordinary least squares multiple regression model predicting 2021 state-level synthetic opioid overdose mortality rates (per 100,000, log-transformed) based on 2021 xylazine seizure rates (per 100,000), adjusting for states’ US Census Region and using robust standard errors (SEs). In an alternate specification, we modeled xylazine seizures as a *percentage* (of all seizure reports of heroin, xylazine, fentanyl-related substances, methamphetamine, and cocaine) instead of as a rate per population, considering that proportion-based measures may better reflect the availability of one substance relative to others, irrespective of state differences in law enforcement operations.^19,24^

## Results

### Rates of Xylazine Seizures and Deaths by Region and Year

Figure 1A depicts xylazine seizure rates and xylazine-involved overdose rates in the US over the years 2018-2021. Xylazine seizure rates increased 172% from 2018-2019, 106% from 2019-2020, and 167% from 2020-2021, while age-adjusted xylazine-involved overdose death rates increased 567% from 2018-2019, 135% from 2019-2020, and 126% from 2020-2021.

**Figure 1.**
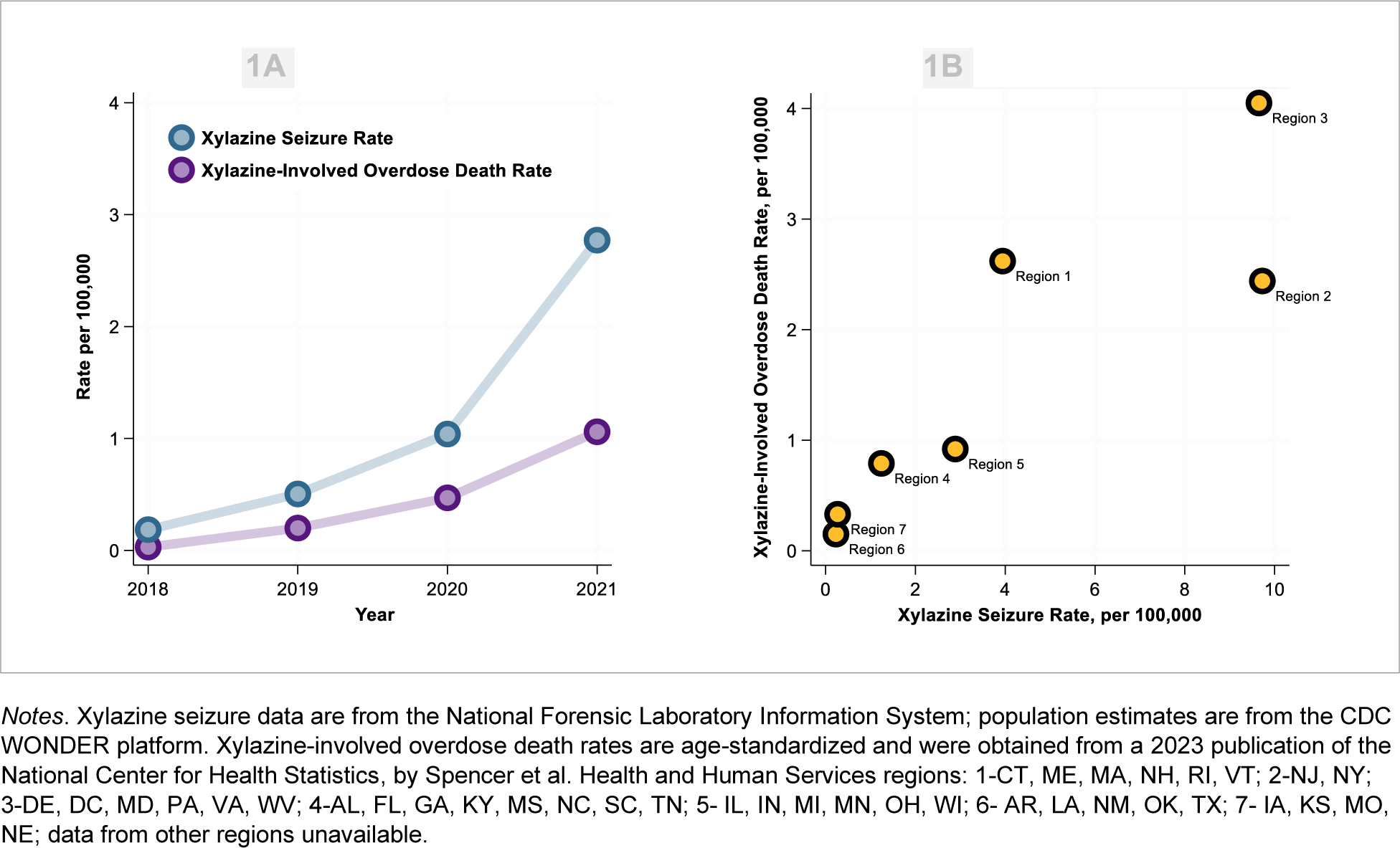
Rates of xylazine seizures and xylazine-involved overdose deaths, per 100,000: a) by year (2018-2021); and b) by Health and Human Services region (2021).

As depicted in Figure 1B, in 2021, HHS regions with higher rates of xylazine seizures were also generally regions with the highest rates of xylazine-involved overdose deaths, while HHS regions with the lowest rates of xylazine seizures also generally exhibited the lowest rates of xylazine-involved overdose deaths. At the same time, xylazine-involved overdose death rates were notably higher in Region 3 (DC, DE, MD, PA, VA, WV) than Region 2 (NJ, NY) even though xylazine seizure rates were similar in the two regions; moreover, while the xylazine seizure rate was more than twice as high in Region 2 (NJ, NY) as Region 1 (CT, MA, ME, NH, RI, VT), xylazine-involved overdose death rates were similar in these two regions.

### State-level Xylazine Seizure Rates

Figure 2 presents xylazine seizure rates by state and year, with states ranked based on their 2022 seizure rate. In 2018, relatively low rates of xylazine seizures were observed across all states, with all states reporting a rate at or below approximately 1 xylazine seizure per 100,000 residents, and 21 states reporting zero seizures. In contrast, in 2022, only three states reported zero xylazine seizures, more than half of states reported a rate above 1 seizure per 100,000 residents, and the highest rates of xylazine seizures (per 100,000 residents) were observed in New Jersey (30.4), Rhode Island (22.7), Maryland (18.9), Virginia (15.5), New Hampshire (13.0), and Ohio (10.9).

**Figure 2.**
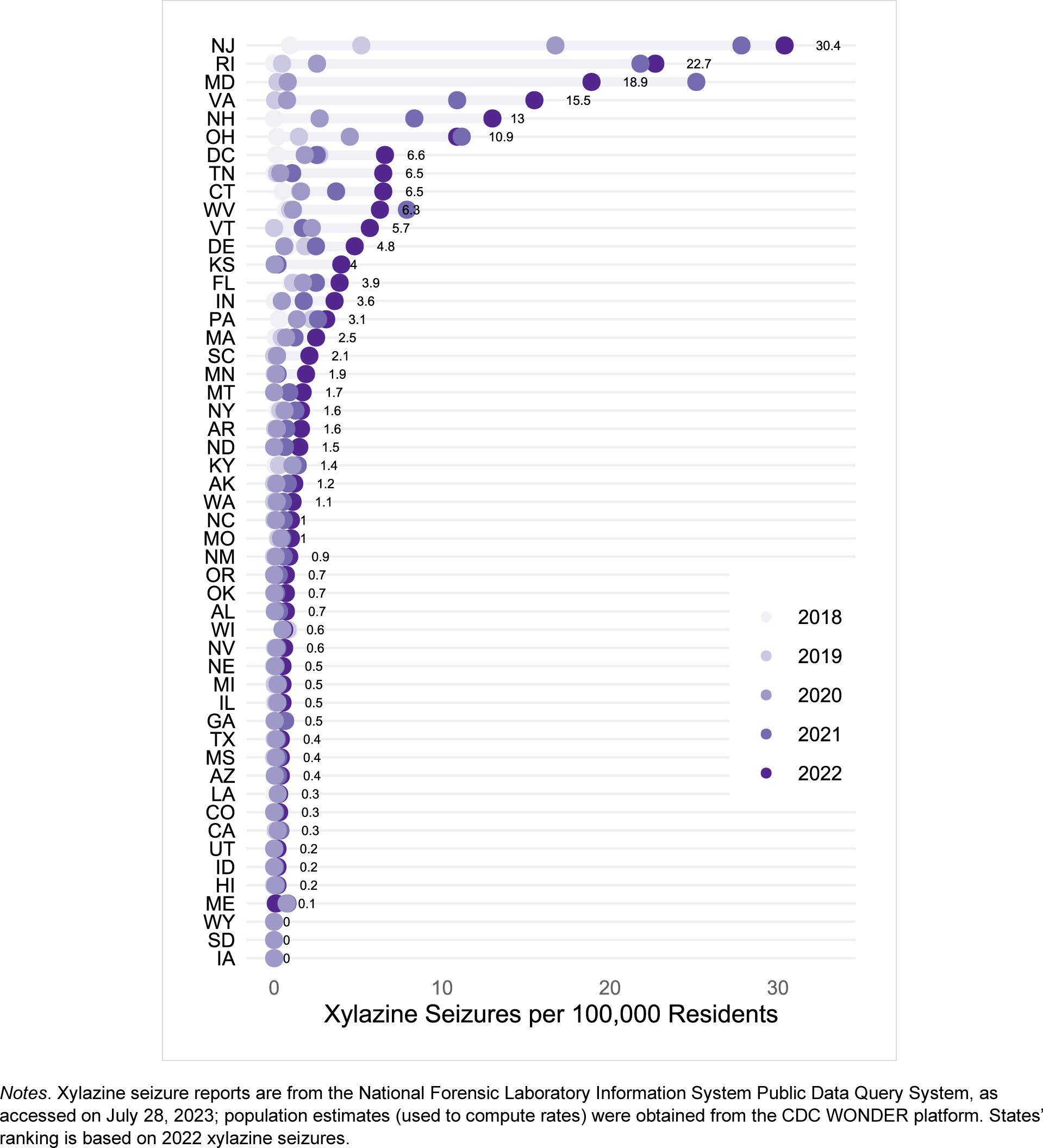
Xylazine law enforcement seizure reports per 100,000 residents, by state and year (2018-2022)

### State-level Xylazine-involved Overdose Death Rates

Table 1 provides numbers and rates of xylazine-involved overdose deaths for each state and year (2019-2022) with data publicly available,^5,8,25–44^ comprising 60 state-year observations across 21 states/DC: 11 states with 2019 data; 15 states with 2020 data; 19 states with 2021 data; and 15 states with 2022 data. Of the states with data publicly available, the highest xylazine-involved overdose death rates (per 100,000 residents) were observed in 2022 in Vermont (10.5) and Connecticut (9.8).

**Table 1.**
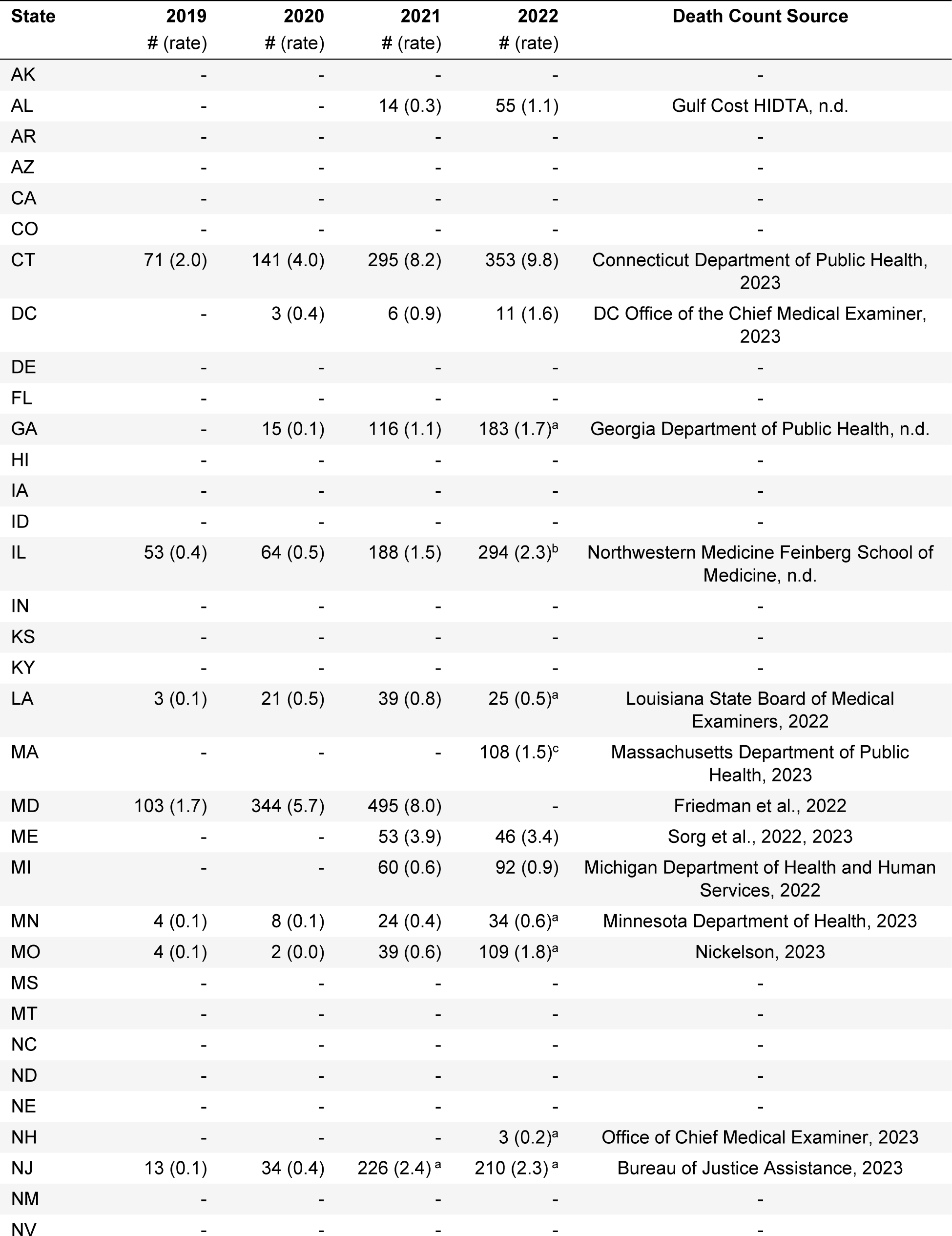
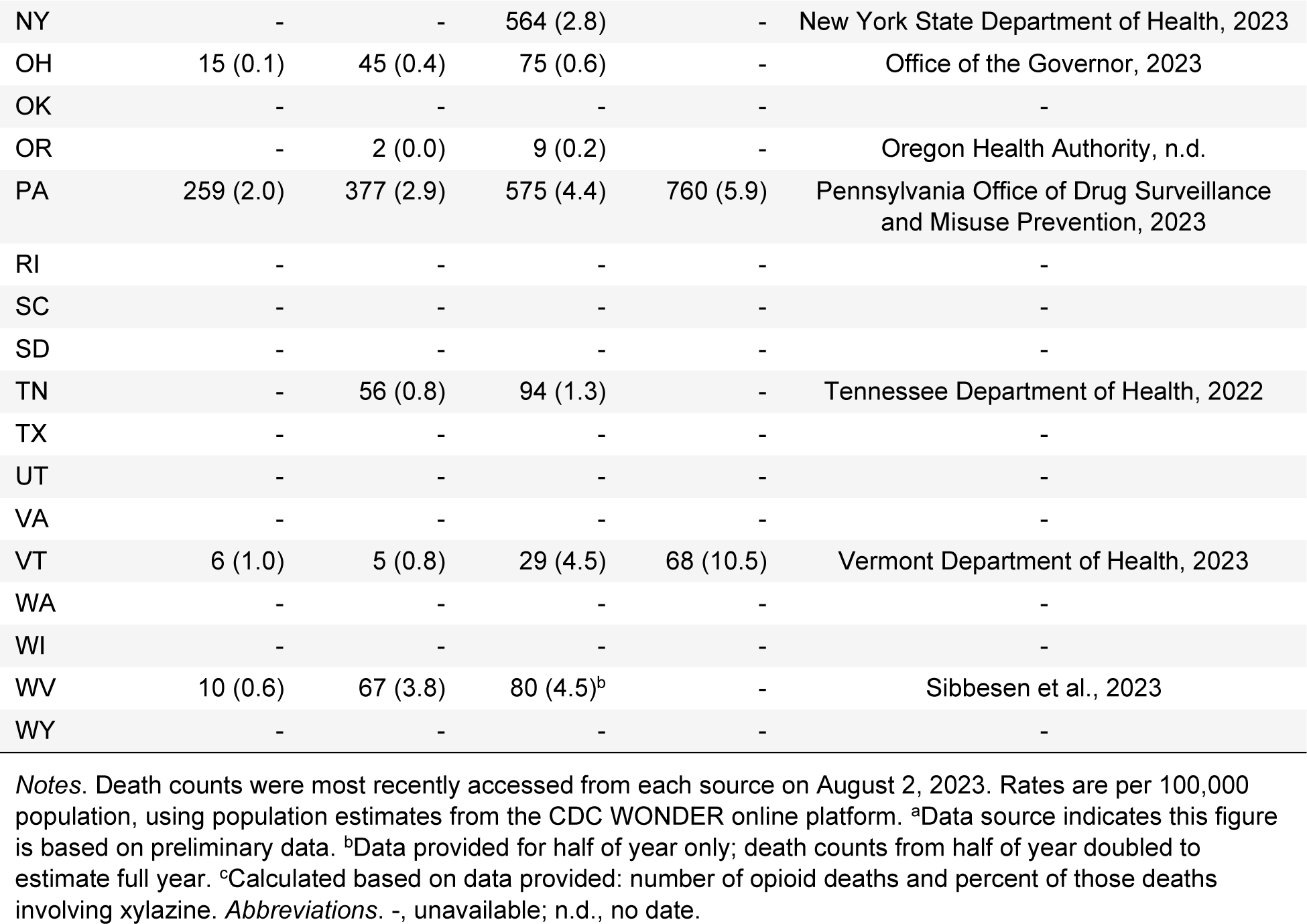
Reported numbers and estimated rates (per 100,000 residents) for overdose deaths involving xylazine, 2019-2022, for states with data available online.

| State | 2019<br># (rate) | 2020<br># (rate) | 2021<br># (rate) | 2022<br># (rate) | Death Count Source |
| --- | --- | --- | --- | --- | --- |
| AK | - | - | - | - | - |
| AL | - | - | 14 (0.3) | 55 (1.1) | Gulf Cost HIDTA, n.d. |
| AR | - | - | - | - | - |
| AZ | - | - | - | - | - |
| CA | - | - | - | - | - |
| CO | - | - | - | - | - |
| CT | 71 (2.0) | 141 (4.0) | 295 (8.2) | 353 (9.8) | Connecticut Department of Public Health, 2023 |
| DC | - | 3 (0.4) | 6 (0.9) | 11 (1.6) | DC Office of the Chief Medical Examiner, 2023 |
| DE | - | - | - | - | - |
| FL | - | - | - | - | - |
| GA | - | 15 (0.1) | 116 (1.1) | 183 (1.7)<sup>a</sup> | Georgia Department of Public Health, n.d. |
| HI | - | - | - | - | - |
| IA | - | - | - | - | - |
| ID | - | - | - | - | - |
| IL | 53 (0.4) | 64 (0.5) | 188 (1.5) | 294 (2.3)<sup>b</sup> | Northwestern Medicine Feinberg School of Medicine, n.d. |
| IN | - | - | - | - | - |
| KS | - | - | - | - | - |
| KY | - | - | - | - | - |
| LA | 3 (0.1) | 21 (0.5) | 39 (0.8) | 25 (0.5)<sup>a</sup> | Louisiana State Board of Medical Examiners, 2022 |
| MA | - | - | - | 108 (1.5)<sup>c</sup> | Massachusetts Department of Public Health, 2023 |
| MD | 103 (1.7) | 344 (5.7) | 495 (8.0) | - | Friedman et al., 2022 |
| ME | - | - | 53 (3.9) | 46 (3.4) | Sorg et al., 2022, 2023 |
| MI | - | - | 60 (0.6) | 92 (0.9) | Michigan Department of Health and Human Services, 2022 |
| MN | 4 (0.1) | 8 (0.1) | 24 (0.4) | 34 (0.6)<sup>a</sup> | Minnesota Department of Health, 2023 |
| MO | 4 (0.1) | 2 (0.0) | 39 (0.6) | 109 (1.8)<sup>a</sup> | Nickelson, 2023 |
| MS | - | - | - | - | - |
| MT | - | - | - | - | - |
| NC | - | - | - | - | - |
| ND | - | - | - | - | - |
| NE | - | - | - | - | - |
| NH | - | - | - | 3 (0.2)<sup>a</sup> | Office of Chief Medical Examiner, 2023 |
| NJ | 13 (0.1) | 34 (0.4) | 226 (2.4)<sup>a</sup> | 210 (2.3)<sup>a</sup> | Bureau of Justice Assistance, 2023 |
| NM | - | - | - | - | - |
| NV | - | - | - | - | - |
| NY | - | - | 564 (2.8) | - | New York State Department of Health, 2023 |
| OH | 15 (0.1) | 45 (0.4) | 75 (0.6) | - | Office of the Governor, 2023 |
| OK | - | - | - | - | - |
| OR | - | 2 (0.0) | 9 (0.2) | - | Oregon Health Authority, n.d. |
| PA | 259 (2.0) | 377 (2.9) | 575 (4.4) | 760 (5.9) | Pennsylvania Office of Drug Surveillance and Misuse Prevention, 2023 |
| RI | - | - | - | - | - |
| SC | - | - | - | - | - |
| SD | - | - | - | - | - |
| TN | - | 56 (0.8) | 94 (1.3) | - | Tennessee Department of Health, 2022 |
| TX | - | - | - | - | - |
| UT | - | - | - | - | - |
| VA | - | - | - | - | - |
| VT | 6 (1.0) | 5 (0.8) | 29 (4.5) | 68 (10.5) | Vermont Department of Health, 2023 |
| WA | - | - | - | - | - |
| WI | - | - | - | - | - |
| WV | 10 (0.6) | 67 (3.8) | 80 (4.5)<sup>b</sup> | - | Sibbesen et al., 2023 |
| WY | - | - | - | - | - |

*Notes.* Death counts were most recently accessed from each source on August 2, 2023. Rates are per 100,000 population, using population estimates from the CDC WONDER online platform. <sup>a</sup>Data source indicates this figure is based on preliminary data. <sup>b</sup>Data provided for half of year only; death counts from half of year doubled to estimate full year. <sup>c</sup>Calculated based on data provided: number of opioid deaths and percent of those deaths involving xylazine. *Abbreviations.* -, unavailable; n.d., no date.

### State Rates of Xylazine Seizures and Xylazine Overdose Deaths

In Figure 3, the magenta lines represent xylazine seizures per 100,000 residents, for each state and DC, 2019-2022, while the blue bars mark xylazine-involved overdose deaths per 100,000 residents for each state and year with data available. In states such as Pennsylvania and Connecticut, visual trends in xylazine-involved overdose death rates appeared to mirror trends in xylazine seizure rates. In contrast, in states such as New Jersey and Ohio, xylazine seizure rates appeared to increase notably more rapidly than xylazine-involved overdose death rates, while in Vermont, rates of xylazine-involved overdose deaths appeared to increase more rapidly than rates of xylazine seizures. Finally, the state with the highest rate of xylazine seizures (New Jersey) reported relatively low rates of xylazine-involved overdose deaths, especially compared to Vermont and Connecticut, states with lower levels of xylazine seizures but higher xylazine overdose death rates.

**Figure 3.**
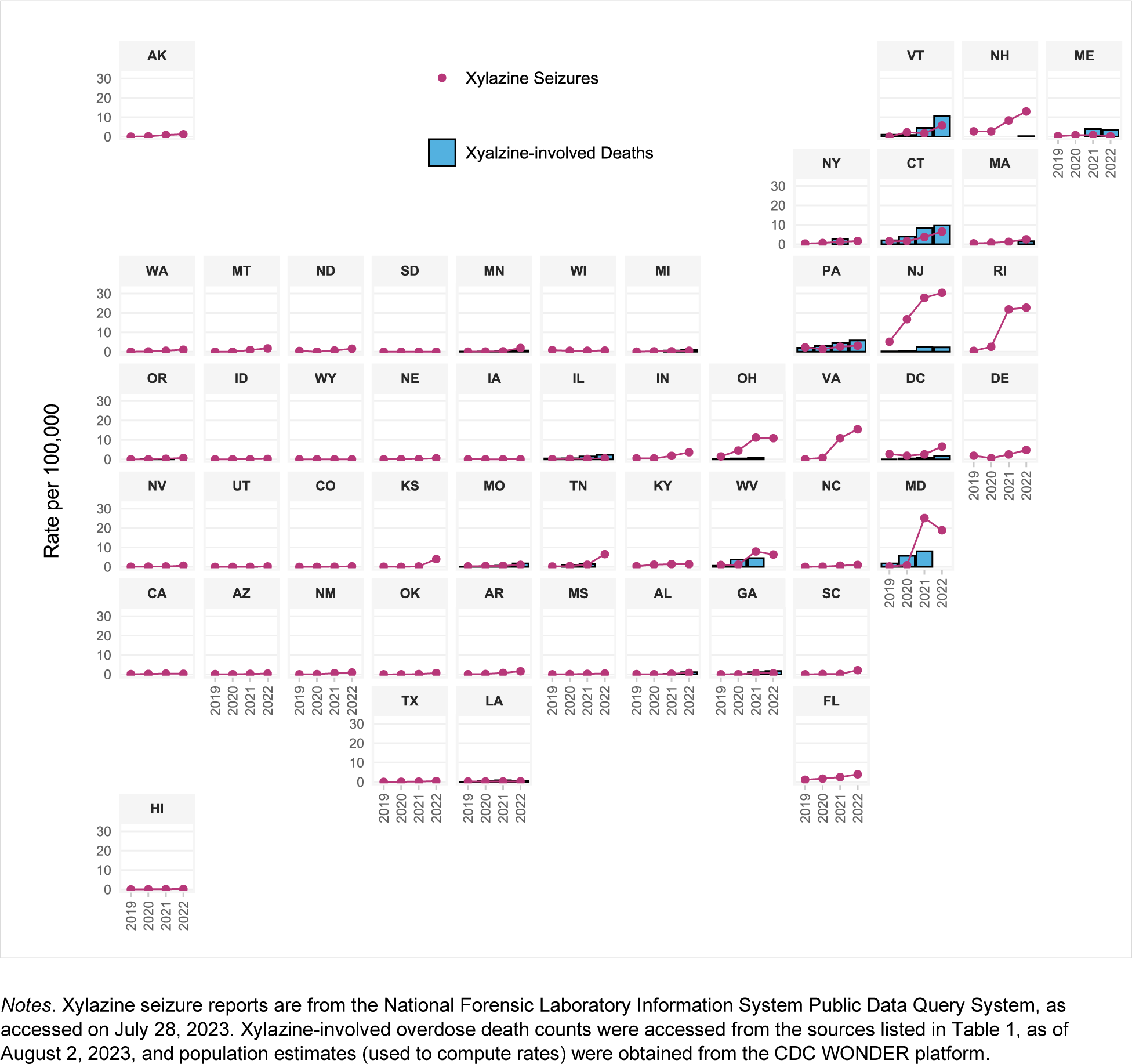
Xylazine law enforcement seizure reports and xylazine-involved overdose deaths, per 100,000 residents, by state and year (2019-2022)

### Association between States’ Xylazine Seizures and Synthetic Opioid Overdose Deaths

In the regression model predicting states’ (*n*=51) 2021 synthetic opioid overdose mortality rates (log-transformed) by xylazine seizure rates, adjusting for US Census Region, each additional xylazine seizure per 100,000 residents was associated with a 2% higher synthetic opioid overdose mortality rate (*b*=0.02, robust se=0.01; [exp(0.02)-1]*100%=2%; *p*=0.049). In the alternate specification of the regression model, the percentage of seizures involving xylazine (instead of the rate per 100,000) was also significantly positively associated with synthetic opioid overdose death rates (*b*=0.05, robust se=0.01; *p*=0.001).

## Limitations

Although the NFLIS reports a 98% participation rate from US forensic laboratories, not every drug seized is analyzed,^22^ and since NFLIS does not represent a random sample of the US illicit drug supply,^45^ it is unclear to what extent drug seizure samples are representative of overall street drug availability. Drug seizure counts from the publicly-available NFLIS also lack information about the weights/dosage equivalents of the drugs seized or the combinations of substances within a single drug seizure,^22^ and different forensic laboratories employ different procedures for testing and reporting.^45^ In the present study, the state-level data on xylazine-involved overdose deaths were located via an online search, resulting in death counts obtained from individual states which may employ different procedures for death investigations, toxicology, death certificate reporting,^2^ and presentation of data on xylazine overdose deaths (e.g., whether including deaths of all intents or only accidental/undetermined intent, presenting all xylazine-positive deaths or only deaths where xylazine was identified as a contributing cause, reporting xylazine-involved deaths or xylazine-fentanyl deaths). Differences in rates of reported xylazine-involved overdose deaths between states, regions, or years may reflect regional and time differences in the extent to which local jurisdictions test for xylazine in post-mortem toxicology.^2^ Finally, in the present study, xylazine-involved overdose death data were located for a limited number of states, precluding statistical examinations of associations between xylazine seizures and xylazine overdose deaths. Moreover, the analysis of the relationship between xylazine seizures and synthetic opioid overdose deaths was cross-sectional and represented a test of association rather than causality.

## Discussion

Results of this study highlight xylazine’s increasing involvement in law enforcement drug seizures and overdose deaths in the US. Rates of xylazine law enforcement seizure reports and xylazine-involved overdose deaths more than doubled each year from 2018-2021, although it is unclear to what extent these increases may reflect expanded testing for xylazine. Study results emphasize the need for more state-level (and community-level) data on xylazine-involved overdose deaths to inform local area overdose response initiatives. In the present study, we were unable to locate yearly xylazine-involved overdose death counts for 30 states, including several states (e.g., VA, DE, RI) bordering high-xylazine mortality states.

Expanding and standardizing postmortem testing and reporting procedures across US jurisdictions has the potential to support more uniformly available and complete data on xylazine-involved overdose deaths.^46,47^ Additionally, since data from drug-checking programs^48–50^ or healthcare settings^51–52^ are currently limited in geographic reach, law enforcement drug seizure data offer a potential gauge of community-level xylazine risk. In the present study, the limited available xylazine overdose death data precluded a formal examination of the association between xylazine seizures and xylazine-involved overdose deaths. Nonetheless, in general, regions and states with higher rates of xylazine seizures also appeared to experience higher rates of xylazine-involved overdose deaths, providing initial support for xylazine seizure rates as potential proxies of xylazine’s presence in illicit drug supplies. At the same time, several differences were observed between state patterns in xylazine seizures and in xylazine-involved overdose deaths; for example, in New Jersey, xylazine seizure rates increased notably more rapidly and far exceeded rates in states such as Vermont and Connecticut, yet xylazine-involved overdose death rates were lower in New Jersey than in Vermont and Connecticut. This example may reflect the geographic differences in drug reporting on death certificates^53^ or the limitations of a drug seizure measure that does not incorporate weight/dosage/drug combinations or differences in law enforcement operations.^17,19^ This finding may also highlight the role of factors beyond xylazine availability that shape state levels of xylazine-involved overdose mortality (e.g., fentanyl/analog availability, socioeconomic contexts, treatment and healthcare services, policies).

In the present study, states’ 2021 xylazine seizure rates were positively associated with 2021 synthetic opioid (e.g., fentanyl) overdose mortality rates, even after adjusting for region. Causal mechanisms were not examined in this preliminary analysis, yet it may be possible that a higher prevalence of xylazine in fentanyl supplies increases fatalities due to xylazine’s effects alone^54^ or in combination with fentanyl,^50^ exacerbated by the lack of a xylazine-reversal agent.^46^ Fatal overdoses involving xylazine nearly always include fentanyl,^2^ and illicitly-manufactured fentanyl has a longer history and notably greater prevalence in eastern states than in the West,^55^ yet the association between xylazine seizures and synthetic opioid overdose mortality rates was observed even after adjusting for states’ Census Region in an attempt to account for this East/West difference.

Overall, study results were consistent with prior studies identifying greater xylazine involvement in overdose deaths in the eastern US,^2^ especially in the Northeast.^3,7^ Adding to the list of states identified as hotspots for xylazine overdose (Maryland,^7^ Connecticut^7^), study results suggest that Vermont has also experienced some of the highest recent reported rates of xylazine-involved overdose deaths. Prior studies have identified Philadelphia, Pennsylvania as the epicenter of xylazine-involved overdose deaths,^1,56^ and in the present study, Pennsylvania reported the highest absolute *number* of xylazine deaths (of all states with available data), yet *rates* of xylazine-involved overdose deaths were higher in several other states (e.g., in 2022: Connecticut, 9.8 per 100,000; Vermont, 10.5) than in Pennsylvania (in 2022: 5.9 per 100,000). At the same time, some Pennsylvania counties evidence high levels of underreporting of drug involvement on death certificates,^53,57^ suggesting that Pennsylvania’s xylazine-involved overdose death rates may be underestimates.

In communities with xylazine prevalent in illicit drug supplies, overdose prevention efforts may be strengthened via the evaluation, optimization, and incorporation of xylazine-specific harm reduction strategies. Recently-developed xylazine test strips represent one such potential harm reduction strategy^58^; research in this area is still nascent,^56,59^ especially relative to research on fentanyl test strips,^60^ and qualitative data suggest that some individuals who use drugs wish to avoid xylazine^56^ while others seek xylazine-adulterated fentanyl for a longer-duration effect.^8^ In addition to test strips, integrating xylazine information in overdose prevention training may better equip individuals who use drugs and other lay and professional overdose responders.^61^ The opioid reversal medication naloxone can counteract the respiratory depression caused by the opioids that commonly accompany xylazine but cannot address the effects of xylazine itself (since xylazine is not an opioid^46^), so lay responders are often advised to not only administer naloxone but also call emergency medical services and provide rescue breathing,^62^ and medical professionals may need to provide additional interventions specific to xylazine effects (e.g., hypotension).^1,46^ While arguably not directly related to overdose, wound care also represents a harm reduction strategy particularly relevant for individuals who use drugs containing xylazine, due to the severe skin ulcerations often accompanying use.^61–63^ These skin ulcerations, as well as the withdrawal symptoms associated with xylazine dependence, also complicate the provision of effective substance use disorder treatment,^1,61^ necessitating additional research and evaluation to optimize treatment care protocols.^47^

## Conclusion

Although xylazine is not currently one of the top drugs contributing to overdose deaths in the US overall, xylazine involvement in overdose deaths is relatively high within certain regions/states/communities,^7,64^ rapidly increasing, and likely underestimated due to limited testing.^2,46^ Moreover, xylazine’s presence in illicit drug supplies is extending across the country, no longer limited to the Northeast.^65,66^ Timely identification of xylazine in local drug supplies and overdoses represents a first step in responding to this “emerging threat,”^1^ followed by the evaluation, optimization, dissemination, and implementation of xylazine-specific harm reduction strategies, including those already identified by frontline workers in regions with a longer history and higher prevalence of xylazine in street drug supplies.^61^

## Data Availability

All data used in the study are publicly available. Drug seizure data are available from https://www.nflis.deadiversion.usdoj.gov/. Synthetic opioid overdose mortality data are available from https://wonder.cdc.gov/mcd.html, and state-level xylazine-involved overdose death data are available from the individual state sources identified in Table 1 of the manuscript and detailed in the reference list.

https://wonder.cdc.gov/mcd.html

